# SARS-CoV-2 incidence and risk factors in a national, community-based prospective cohort of U.S. adults

**DOI:** 10.1101/2021.02.12.21251659

**Authors:** Denis Nash, Madhura S. Rane, Mindy Chang, Sarah Gorrell Kulkarni, Rebecca Zimba, William You, Amanda Berry, Chloe Mirzayi, Shivani Kochhar, Andrew Maroko, McKaylee M. Robertson, Drew A. Westmoreland, Angela M. Parcesepe, Levi Waldron, Christian Grov

## Abstract

**Background:** Epidemiologic risk factors for incident SARS-CoV-2 infection as determined via prospective cohort studies greatly augment and complement information from case-based surveillance and cross-sectional seroprevalence surveys.

**Methods:** We estimated the incidence of SARS-CoV-2 infection and risk factors in a well-characterized, national prospective cohort of 6,738 U.S. adults, enrolled March-August 2020, a subset of whom (n=4,510) underwent repeat serologic testing between May 2020 and January 2021. We examined the crude associations of sociodemographic factors, epidemiologic risk factors, and county-level community transmission with the incidence of seroconversion. In multivariable Poisson models we examined the association of social distancing and a composite score of several epidemiologic risk factors with the rate of seroconversion.

**Findings:** Among the 4,510 individuals with at least one serologic test, 323 (7.3%, 95% confidence interval [CI] 6.5%-8.1%) seroconverted by January 2021. Among 3,422 participants seronegative in May-September 2020 and tested during November 2020-January 2021, we observed 161 seroconversions over 1,646 person-years of follow-up (incidence rate of 9.8 per 100 person-years [95%CI 8.3-11.5]). In adjusted models, participants who reported always or sometimes social distancing with people they knew (IRR_always vs. never_ 0.43, 95%CI 0.21-1.0; IRR_sometimes vs. never_ 0.47, 95%CI 0.22-1.2) and people they did not know (IRR_always vs. never_ 0.64, 95%CI 0.39-1.1; IRR_sometimes vs. never_ 0.60, 95%CI 0.38-0.97) had lower rates of seroconversion. The rate of seroconversion increased across tertiles of the composite score of epidemiologic risk (IRR_medium vs. low_ 1.5, 95%CI 0.92-2.4; IRR_high vs. low_ 3.0, 95%CI 2.0-4.6). Among the 161 observed seroconversions, 28% reported no symptoms of COVID-like illness (i.e., were asymptomatic), and 27% reported a positive SARS-CoV-2 diagnostic test. Ultimately, only 29% reported isolating and 19% were asked about contacts.

**Interpretation:** Modifiable epidemiologic risk factors and poor reach of public health strategies drove SARS-CoV-2 transmission across the U.S during May 2020-January 2021.

**Funding:** U.S. National Institutes of Allergy and Infectious Diseases (NIAID).

## INTRODUCTION

A major challenge of controlling community transmission of SARS-CoV-2 is that the virus’ infectious period allows for onward spread without, or prior to, diagnosis of infection[1,2], including by fully-vaccinated individuals.[3] One national study in the United States (U.S.) prior to the vaccine era estimated that there were 5 undiagnosed infections for every diagnosed case.[2]

While SARS-CoV-2 is understood to be transmitted from person-to-person via airborne and droplet spread, to date, the incidence of SARS-CoV-2 infection and risk factors for incident infection have not been adequately characterized by routine case-based surveillance of SARS-CoV-2 diagnoses or by cross-sectional seroprevalence studies.[4–6] It is critical for prospective studies to investigate COVID-19’s evolving epidemiology and risk factors for SARS-CoV-2 acquisition in communities, including the uptake and impact of non-pharmaceutical interventions (NPIs)[7], and the reach of public health strategies aimed at controlling community transmission, including testing, quarantine, isolation, contact tracing, and vaccination.

Globally, few community-based prospective epidemiologic studies of SARS-CoV-2 incidence and risk factors have been undertaken. One recent global systematic review of observational studies of SARS-CoV-2 that employed serologic or polymerase chain reaction (PCR) testing found 18 prospective studies.[8] Most were focused on healthcare workers or other occupational groups, individuals in congregate settings, evacuees, or cruise ship patrons; none were community-based (i.e., focused on risk factors in communities vs other higher risk populations/settings).[8] Since then, there have been publications from a national community-based prospective cohort in the U.K. [9–11], but to date, there have been no prospective community-based studies with biomarkers in the U.S. Such studies are needed to help inform aspects of implementation of the public health response and policies, both to the current pandemic and future ones.

In late March 2020, we launched the prospective Communities, Households and SARS-CoV-2 Epidemiology (CHASING) COVID Cohort.[12] We describe the incidence of SARS-CoV-2 infection and risk factors for SARS-CoV-2 seroconversion during May 2020-January 2021, as well as the reach and uptake of public health strategies aimed at controlling community spread among those who seroconverted.

## METHODS

### Recruitment

We used internet-based strategies[13,14] to recruit a geographically and socio-demographically diverse cohort of adults into longitudinal follow-up with at-home specimen collection. To be eligible for inclusion in the cohort, individuals had to: 1) reside in the U.S. or a U.S. territory; 2) be ≥18 years old; 3) provide a valid email address for follow-up; and 4) demonstrate early engagement in study activities (provision of a baseline specimen or completion of >1 recruitment/enrollment visit). Details of the study design and recruitment procedures are described elsewhere.[15] The full cohort includes participants from all 50 U.S. states, the District of Columbia, Puerto Rico, and Guam (Supplemental Figure S1). Of the 6,738 participants in the full cohort, 4,510 (67%) had at least one serologic test and comprised the study population for this analysis (Table S1).

### Data collection

Cohort recruitment and enrollment visits were completed between March 28-August 21, 2020, during which multiple rounds of interviews took place. Demographic and COVID-19 related risk factors were collected at baseline. From three follow-up interviews between August and November 2020, we obtained repeated measurements of COVID-19 symptoms, laboratory testing (PCR or serologic), hospitalizations, use of NPIs such as mask use and social distancing, public health strategies such as quarantine, isolation, and contact tracing encounters, and other time-varying factors.

During May-September 2020 (Period 1) and November 2020-January 2021 (Period 2), participants were invited to complete serologic testing using an at-home self-collected dried blood spot (DBS) specimen collection kit. DBS cards were sent from and returned to the study laboratory (Molecular Testing Laboratories [MTL], Vancouver, Washington) via the U.S. Postal Service using a self-addressed, stamped envelope containing a biohazard bag™.

All DBS specimens were tested by the study laboratory for total antibodies (Total Ab) using the Bio-Rad Platelia test for IgA, IgM, and IgG which targets the SARS-CoV-2 nucleocapsid protein (manufacturer sensitivity 98.0%, specificity 99.3%).[16] Other studies have independently validated this assay and found average sensitivity and specificity of 91.7% and 98.8%, respectively.[17–19] This assay was also validated for use with DBS by the study laboratory, which found 100% sensitivity and 100% specificity (MTL, personal communication).

### Outcomes

#### Cumulative incidence of SARS-CoV-2 infection

Among participants who underwent serologic testing, we estimated the serology-based cumulative incidence of SARS-CoV-2 as the proportion of individuals with a positive Total Ab test in either of the two time periods (i.e., number of participants ever positive) divided by the total number of persons with one or more Total Ab tests (i.e., number of participants ever tested). We adjusted cumulative incidence estimates for laboratory test error, assuming a sensitivity of 91.7% and a specificity of 98.8%[17–19] using the following formula[20]:

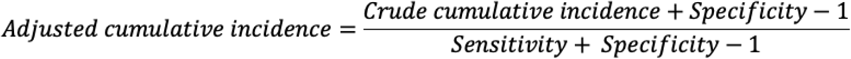

#### Observed SARS-CoV-2 seroconversion

Among those individuals with two Total Ab tests, an observed seroconversion was defined as a negative Total Ab test in Period 1 followed by a positive Total Ab test in Period 2. We estimated person-years of follow-up using the collection dates for each specimen in Periods 1 and 2. When the collection date was missing, we used the date the laboratory received the sample. The seroconversion date was assigned as the midpoint between the first and second specimen collection dates for person-time calculations.

### Exposures

#### Individual-level COVID-19 risk factors

We examined epidemiologic risk factors for SARS-CoV-2 individually and as part of a composite score. We considered the following risk behaviors and factors reported by participants prior to specimen collection: Household factors (household crowding [4 or more people living in a household], having a child in the household, and having a confirmed COVID-19 case in a household member prior to participant testing positive), spending time in public places (attending mass gatherings such as protests, spending time in an indoor restaurant or bar, spending time at an outdoor restaurant or bar, visiting places of worship, or visiting public parks or pools), mask use indoors (for grocery shopping, visiting non-household members, at work, and in salons or gyms), mask use outdoors, gathering in groups with more than 10 people (indoors only, outdoors only, and indoors and outdoors), travel during the pandemic (recent air travel and public transit use), and individual-level factors that may increase the risk of severe COVID-19 (substance use, binge drinking, and medically diagnosed comorbidities).

#### Global social distancing assessment

While social distancing in specific scenarios is addressed in some of the above individual risk factors (such as spending time in various public places), we were specifically interested in the association between social distancing in general and incident SARS-CoV-2 infection. We asked two global questions on social distancing, which were “In the past month, how often have you practiced social distancing with: a) *people you know* and b) *people you do not know*,” with possible response options of *Always, Sometimes, or Never*. These assessments were not included in the calculation of the composite risk score.

#### Composite score of COVID-19 risk factors

We computed a composite COVID-19 risk score as many of the above COVID-19 risk factors may be highly correlated. We applied Least Absolute Shrinkage Selection Operator (LASSO) regression to select the set of risk factors which best predicted seroconversion.[21] Scores were assigned to each participant based on whether they engaged in the risk factors selected by the LASSO regression model and were normalized between 0 and 100. High scores indicate engagement in high-risk activities. Details are in the Statistical Appendix. The composite score was divided into tertiles (low, medium, high) for analysis.

### Statistical analysis

Cumulative incidence estimates were stratified by baseline characteristics and epidemiologic risk factors. Crude and adjusted Incidence Rate Ratios (IRRs) of seroincidence and associated 95% confidence intervals (CIs) were estimated using Poisson regression. We examined crude seroconversion rates by sociodemographic characteristics and each risk factor. Finally, we separately modeled three exposure variables: 1) social distancing with “people you know” (Yes/No); 2) social distancing with “people you don’t know” (Yes/No); and 3) the composite COVID-19 risk score(high/medium/low). Two multivariable models were constructed for each exposure variable, including models that adjusted for age, gender, race/ethnicity and comorbidities (Model 1); and a model that further controlled for changes in community-level COVID-19 transmission (Model 2). See Statistical Appendix for details. All data were cleaned and analyzed in R (version 4.0.3) and SAS (V9.4).

### Ethical Approval

The study protocol was approved by the Institutional Review Board at the City University of New York (CUNY).

## RESULTS

### Sample characteristics

The characteristics of the study sample are shown in Table 1. A total of 4,232 persons underwent serologic testing in Period 1, and 3,883 were tested in Period 2 (Table S1). Of the 4,510 participants who tested at least once, 3,605 (80%) tested at both time points (Table 1). Differences between those participants testing in Period 1 and Period 2 were negligible (Table S1). The median time between specimen collection dates for participants providing specimens for both serologic tests was 190 days (IQR 152-201) (Figure S2).

**Table 1.** Baseline characteristics of CHASING COVID Cohort Study participants who provided a dried blood spot sample for antibody testing

|  | Participants with one or more serologic tests |  |
| --- | --- | --- |
|  | N | % |
| **Total** | 4,510 | 100.00 |
| Two serologic tests | 3,605 | 79.93 |
| **Age** |  |  |
| Median (IQR) | 41 (31, 55) |  |
| 18-29 | 876 | 19.42 |
| 30-39 | 1,253 | 27.78 |
| 40-49 | 858 | 19.02 |
| 50-59 | 658 | 14.59 |
| 60+ | 865 | 19.18 |
| **Gender** |  |  |
| Male | 2,018 | 44.75 |
| Female | 2,360 | 52.33 |
| Non-Binary/Transgender | 132 | 2.93 |
| **Race/Ethnicity** |  |  |
| Non-Hispanic White | 2,966 | 65.76 |
| Hispanic | 679 | 15.06 |
| Non-Hispanic Black | 378 | 8.38 |
| Asian/PI | 304 | 6.74 |
| Other | 168 | 3.73 |
| Missing | 15 | 0.33 |
| **Education** |  |  |
| Less than high school | 54 | 1.20 |
| High school graduate | 403 | 8.94 |
| Some college | 1,151 | 25.52 |
| College graduate | 2,902 | 64.35 |
| **Employment** |  |  |
| Employed | 2,774 | 61.51 |
| Out of work | 544 | 12.06 |
| Homemaker | 242 | 5.37 |
| Student | 367 | 8.14 |
| Retired | 583 | 12.93 |
| **Household income** |  |  |
| Less than \$35,000 | 1,186 | 26.30 |
| \$35-49,999 | 496 | 11.00 |
| \$50-69,999 | 643 | 14.26 |
| \$70-99,999 | 741 | 16.43 |
| \$100,000+ | 1,329 | 29.47 |
| Don't know | 112 | 2.48 |
| Missing | 3 | 0.07 |
| **Setting** |  |  |
| Urban | 1,911 | 42.37 |
| Suburban | 1,186 | 26.30 |
| Rural | 1,412 | 31.31 |
| Missing | 1 | 0.02 |
| **Geographic region** |  |  |
| Northeast | 1,320 | 29.27 |
| Midwest | 805 | 17.85 |
| South | 1,269 | 28.14 |
| West | 1,111 | 24.63 |
| US Territories | 5 | 0.11 |
| **Healthcare worker** |  |  |
| No | 4,048 | 89.76 |
| Yes | 425 | 9.42 |
| Don't know | 37 | 0.82 |
| **Essential worker \*** |  |  |
| No | 3,170 | 70.29 |
| Yes | 1,340 | 29.71 |
| **Higher risk for severe COVID \*\*** |  |  |
| No | 2,097 | 46.50 |
| Yes | 2,413 | 53.50 |

\* Combined from three follow-up interviews between August and November 2020
\*\* >60 years old, or reported co-morbidity, or current smoker. Comorbidity was defined as having history of heart attack, depression, angina, immunosuppression, type 2 diabetes, high blood pressure, cancer, asthma, COPD, chronic kidney disease, and/or HIV/AIDS

### Cumulative incidence of SARS-CoV-2 as of January 31, 2021

We observed 323 seropositives among the 4,510 participants who tested at least once during follow-up, for an overall crude and adjusted serology-based estimates of cumulative incidence of 7.3% (95% confidence interval [CI]: 6.5%-8.1%) and 6.7% (95% CI 5.9%-7.6%), respectively (Table S2).

### SARS-CoV-2 seroincidence, May 2020-January 2021

There were 3,422 participants who were seronegative in Period 1 with a subsequent serologic test in Period 2 who were followed prospectively for the outcome of SARS-CoV-2 seroconversion. There were 161 observed seroconversions over 1,646 person years of follow-up, for an overall incidence rate of 9.8 per 100 person-years (95% CI 8.3-11.5) (Table 2). The rate of incident SARS-CoV-2 infection was lower for females compared to males (IRR=0.69, 95% CI:0.50, 0.94), and higher for Hispanics (IRR=2.09, 95% CI 1.41-3.05) and non-Hispanic Blacks (IRR=1.69, 95% CI 0.96-2.82) compared with non-Hispanic Whites. Essential workers had higher incidence than non-essential workers (IRR=1.65, 95% CI 1.10-2.26). Incidence rates were higher among those in rural versus urban areas (IRR=1.29, 95% CI 0.89-1.29), and among those in the Midwest (IRR=1.59, 95% CI 0.98-2.56), the South (IRR=1.67, 95% CI 1.08-2.59), and the West (IRR=1.32, 95% CI 0.83-2.11) compared to the Northeast.

**Table 2:**
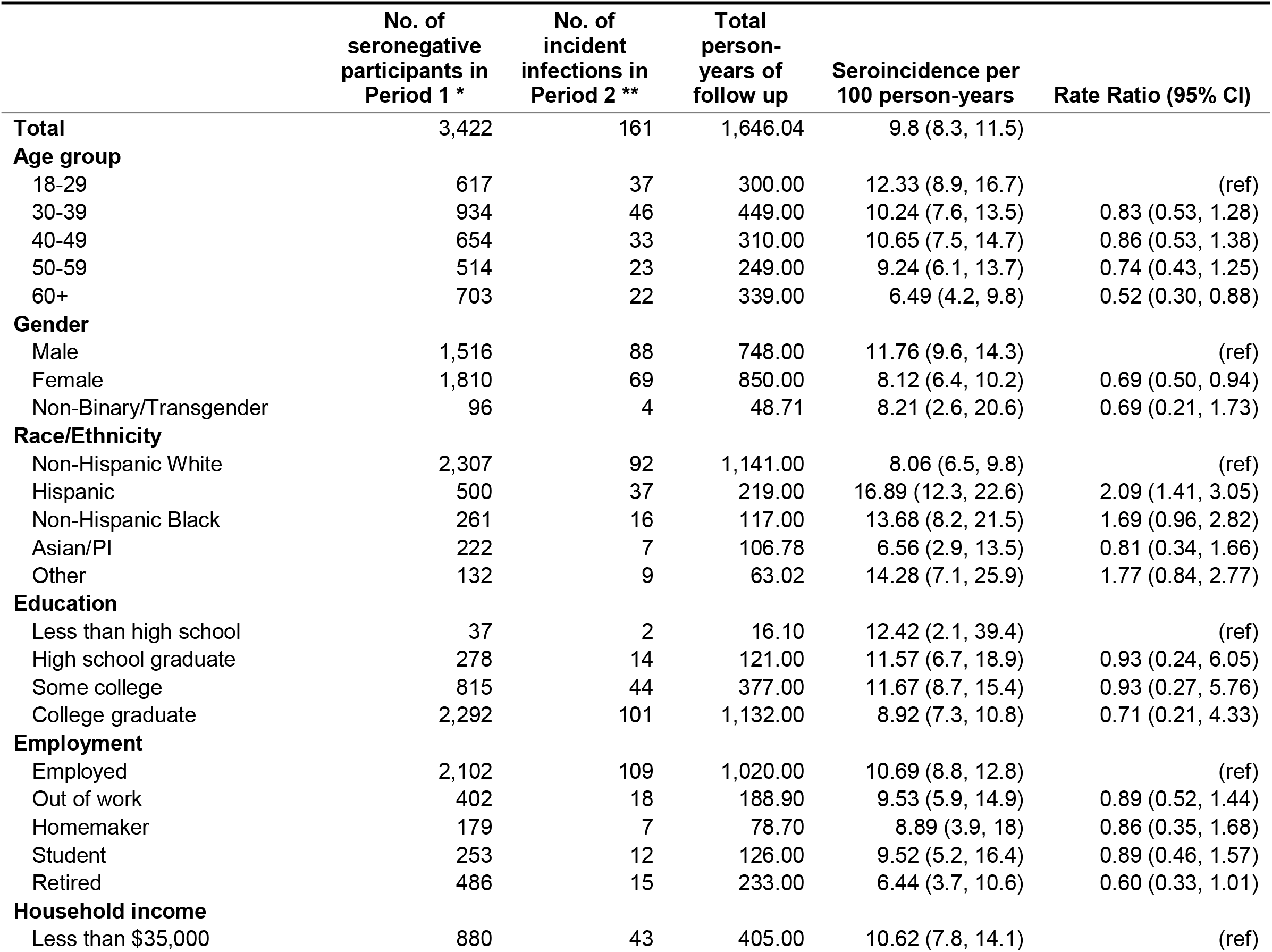
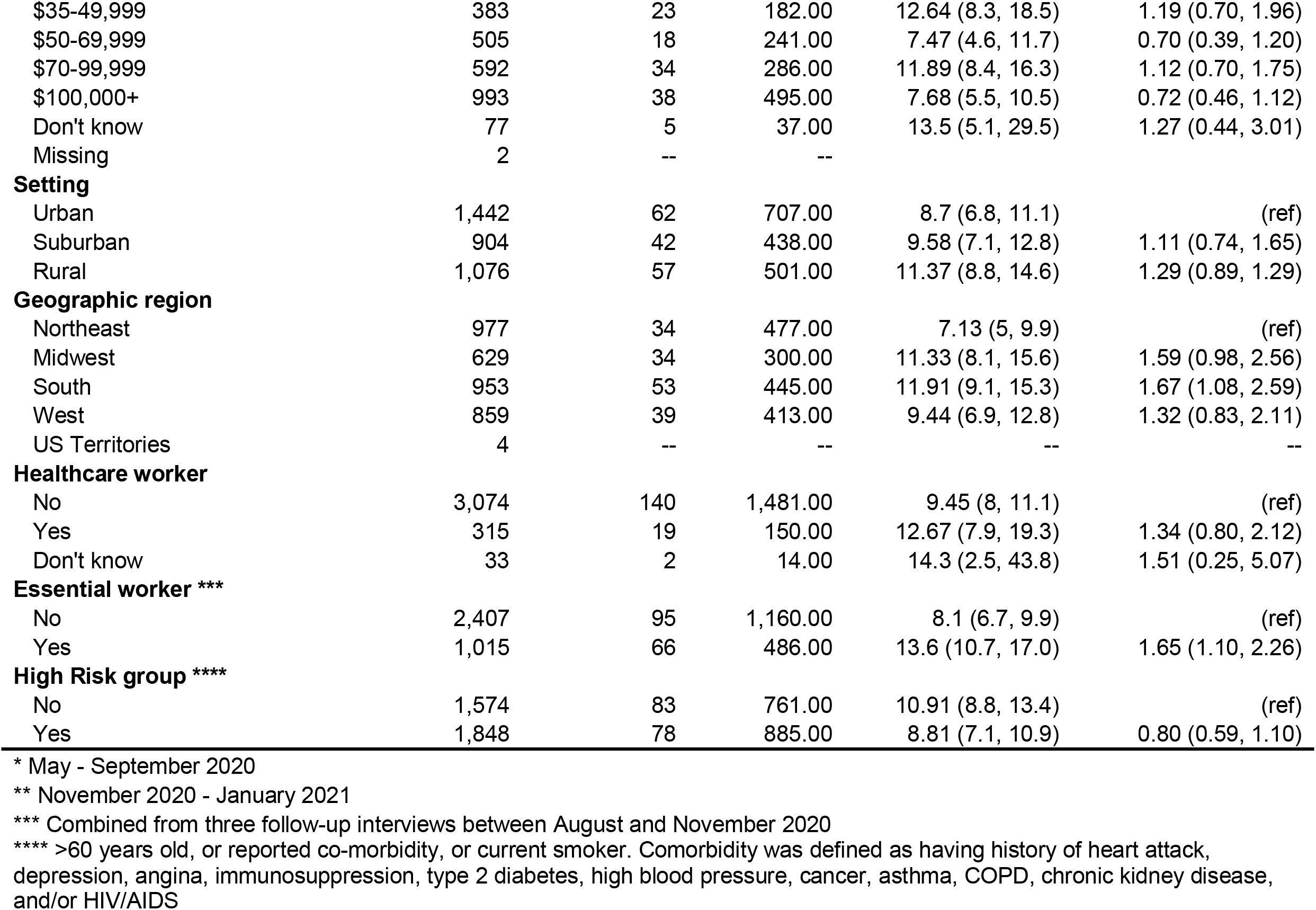
Crude associations of COVID-19 sociodemographic factors with seroincidence, May 2020-January 2021

Table 3 shows the seroincidence and crude incidence rate ratios by epidemiologic risk factors that were present prior to or between serologic tests. There was higher incidence among those who dined indoors at restaurants or bars (IRR=1.93, 95% CI 1.39-2.70); those who visited a place of worship (IRR=1.92, 95% CI 1.26-2.84); those who wore a mask only sometimes while grocery shopping (IRR=10.57, 95% CI 4.00-30.51); those who visited indoors with people not in their own household while sometimes wearing a mask (IRR=1.94; 95% CI 1.37-3.31) or while never wearing a mask (IRR=2.62; 95% CI 1.50-4.70); those working indoors at a place of employment while never wearing a mask (IRR=2.50, 95% CI 0.98-5.26); those who wore masks only sometimes while attending a salon or gym (IRR=3.23, 95% CI 1.90-5.23); and those who reported traveling by air during August-November 2020 (IRR=1.52, 95% CI 1.05-2.17).

**Table 3:** Crude associations of COVID-19 risk factors with seroincidence, May 2020-January 2021

|  | No. of<br>seronegative<br>participants in<br>Period 1 * | No. of<br>incident<br>infections in<br>Period 2 ** | Total person-years of<br>follow up | Incidence rate per<br>100 person-years<br>(95% CI) | Rate Ratio<br>(95% CI) |
| --- | --- | --- | --- | --- | --- |
| **Total** | 3,422 | 161 | 1,646.04 | 9.8 (8.3, 11.5) |  |
| **Household factors** |  |  |  |  |  |
| Household crowding | 822 | 43 | 378.88 | 11.4 (8.3, 15.4) | 1.21 (0.85, 1.71) |
| No household crowding | 2,600 | 118 | 1,267.16 | 9.3 (7.7, 11.2) | ref |
| Child in household | 653 | 28 | 296.07 | 9.5 (6.5, 13.8) | 0.95 (0.62, 1.42) |
| No child in household | 2,769 | 133 | 1,349.97 | 9.9 (8.3, 11.7) | ref |
| Confirmed case in household member | 23 | 12 | 102.28^ | 11.7 (5.2, 26.6) | 15.7 (8.71, 28.27) |
| No confirmed case in household member | 3,399 | 149 | 19,938.30^ | 0.7 (0.6, 0.9) | ref |
| **Social distancing** |  |  |  |  |  |
| *Social distancing with people you know* |  |  |  |  |  |
| Always | 2,616 | 113 | 1,258.00 | 8.9 (7.4, 10.7) | 0.29 (0.14, 0.69) |
| Sometimes | 660 | 34 | 319.00 | 10.6 (7.6, 14.7) | 0.35 (0.16, 0.86) |
| Never | 53 | 7 | 23.30 | 30.0 (13.8, 52.4) | ref |
| Not Applicable | 53 | 4 | 24.50 | 16.3 (5.3, 37.5) | 0.53 (0.13, 1.83) |
| *Social distancing with people you do not know* |  |  |  |  |  |
| Always | 1,137 | 51 | 543.00 | 9.4 (7.1, 12.2) | 0.54 (0.33, 0.89) |
| Sometimes | 1,787 | 79 | 868.00 | 9.1 (7.3, 11.3) | 0.52 (0.33, 0.84) |
| Never | 300 | 24 | 139.00 | 17.3 (11.5, 24.8) | ref |
| Not Applicable | 158 | 4 | 74.50 | 5.4 (1.7, 13.9) | 0.31 (0.09, 0.83) |
| **Spent time in public places** |  |  |  |  |  |
| Attended mass gathering(s) | 350 | 18 | 174.89 | 10.3 (6.4, 16.5) | 1.05 (0.63, 1.69) |
| Did not attend mass gathering(s) | 3,072 | 143 | 1,471.15 | 9.7 (8.2, 11.5) | ref |
| Indoor dining/bar | 1,755 | 108 | 843.86 | 12.8 (10.5, 15.5) | 1.93 (1.39, 2.70) |
| No indoor dining/bar | 1,667 | 53 | 802.18 | 6.6 (5.0, 8.7) | ref |
| Outdoor dining/bar | 1,869 | 97 | 924.35 | 10.5 (8.6, 12.9) | 1.18 (0.63, 1.62) |
| No outdoor dining/bar | 1,553 | 64 | 721.70 | 8.9 (6.9, 11.4) | ref |
| Visited place of worship | 359 | 29 | 168.87 | 17.2 (11.7, 25.1) | 1.92 (1.26, 2.84) |
| Did not visit place of worship | 3,063 | 132 | 1,477.17 | 8.9 (7.5, 10.6) | ref |
| Visited public park/public pool | 2,189 | 100 | 1,076.41 | 9.3 (7.6, 11.4) | 0.86 (0.63, 1.19) |
| Did not visit public park/pool | 1,233 | 61 | 569.63 | 10.7 (8.3, 13.9) | ref |
| **Gathered in groups ≥10** |  |  |  |  |  |
| No | 126 | 6 | 63.11 | 9.5 (4.2, 21.6) | ref |
| Indoors only | 231 | 11 | 109.34 | 10.1 (5.5, 18.4) | 1.06 (0.39, 3.09) |
| Outdoors only | 647 | 38 | 318.32 | 11.9 (8.6, 16.6) | 1.26 (0.55, 3.26) |
| Indoors and outdoors | 2,418 | 106 | 1,155.28 | 9.2 (7.6, 11.1) | 0.96 (0.45, 2.42) |
| **Mask Use** |  |  |  |  |  |
| *Mask while grocery shopping* |  |  |  |  |  |
| Did not go grocery shopping | 133 | 6 | 63.43 | 9.5 (4.2, 21.5) | ref |
| Always | 3,084 | 140 | 1,489.12 | 9.4 (7.9, 11.1) | 0.99 (0.46, 2.99) |
| Sometimes | 132 | 12 | 57.22 | 21.0 (11.6, 38.0) | 10.57 (4.00, 30.51) |
| Never | 33 | 0 | 15.11 | 0 | 0 (0, 2.71) |
| *Mask while indoors visiting non-household members* |  |  |  |  |  |
| N/A (Did not visit non-household members indoors) | 669 | 19 | 319.92 | 5.9 (3.8, 9.4) | ref |
| Always | 1,118 | 43 | 546.26 | 7.9 (5.8, 10.7) | 1.32 (0.77, 2.32) |
| Sometimes | 1,131 | 63 | 546.44 | 11.5 (8.9, 14.9) | 1.94 (1.37, 3.31) |
| Never | 463 | 33 | 211.58 | 15.6 (10.9, 22.22) | 2.62 (1.5, 4.7) |
| *Mask while indoors at work* |  |  |  |  |  |
| N/A (Did not attend indoor workplace) | 1,642 | 63 | 789.46 | 8.0 (6.2, 10.3) | ref |
| Always | 1,372 | 71 | 660.60 | 10.7 (8.5, 13.6) | 1.34 (0.95, 1.89) |
| Sometimes | 299 | 18 | 144.08 | 12.5 (7.8, 20.1) | 1.56 (0.90, 2.60) |
| Never | 68 | 6 | 30.05 | 20.0 (8.6, 46.2) | 2.50 (0.98, 5.26) |
| *Mask while at salon/gym* |  |  |  |  |  |
| N/A (Did not attend salon/gym) | 1,567 | 56 | 749.79 | 7.5 (5.7, 9.8) | ref |
| Always | 1,527 | 75 | 743.45 | 10.1 (8.0, 12.7) | 1.31 (0.95, 1.91) |
| Sometimes | 180 | 20 | 82.69 | 24.2 (15.2, 38.5) | 3.23 (1.90, 5.23) |
| Never | 108 | 7 | 48.94 | 14.3 (6.6, 30.8) | 1.91 (0.80, 4.01) |
| *Outdoor mask use* |  |  |  |  |  |
| Mask use outdoors | 1,562 | 67 | 753.27 | 8.9 (7.0, 11.4) | 0.84 (0.67, 1.15) |
| No mask use outdoors | 1,860 | 94 | 892.78 | 10.5 (8.6, 13.0) | ref |
| **Movement during the pandemic** |  |  |  |  |  |
| *Use of public transit* |  |  |  |  |  |
| Avoided or did not use | 2,975 | 138 | 1,434.70 | 9.6 (8.1, 11.4) | 0.88 (0.57, 1.40) |
| Used public transit | 447 | 23 | 211.35 | 10.9 (7.2, 16.6) | ref |
| *Recent air travel (August – November 2020)* |  |  |  |  |  |
| Yes | 582 | 39 | 287.91 | 13.5 (9.8, 18.7) | 1.52 (1.05, 2.17) |
| No | 2,800 | 119 | 1,336.97 | 8.9 (7.4, 10.7) | ref |
| Missing | 40 | 3 | 21.17 | 14.2 (4.4, 46.0) | -- |
| **Other potential risk factors** |  |  |  |  |  |
| Alcohol use *** | 523 | 37 | 249.28 | 14.8 (10.6, 20.7) | 1.67 (1.14, 2.39) |
| Substance use **** | 914 | 45 | 443.40 | 10.1 (7.5, 13.7) | 1.05 (0.73, 1.47) |
| Any comorbidities ^^ | 1,490 | 69 | 714.52 | 9.7 (7.6, 12.3) | 0.97 (0.71, 1.33) |
| **Changes in county-level community transmission** |  |  |  |  |  |
| High (0.327-3.30) | 1,139 | 76 | 536.00 | 14.2 (11.4, 17.5) | ref |
| Medium (0.211-0.327) | 1,140 | 37 | 557.00 | 6.6 (4.8, 9.1) | 0.46 (0.31, 0.69) |
| Low (0-0.211) | 1,139 | 48 | 550.00 | 8.7 (6.5, 11.4) | 0.61 (0.42, 0.88) |
| Missing | 4 | -- | 1.00 |  |  |

\* May - September 2020
\*\* November 2020 - January 2021
\*\*\* Having more than 6 alcoholic drinks on one occasion in last month
\*\*\*\* Used prescription opioids, street opioids, or cannabis in last month
^ Person-time is in person-months
^^ >60 years old, or reported co-morbidity, or current smoker. Comorbidity was defined as having history of heart attack, depression, angina, immunosuppression, type 2 diabetes, high blood pressure, cancer, asthma, COPD, chronic kidney disease, and/or HIV/AIDS

### Poisson models of SARS-CoV-2 seroconversion for three different exposures of interest, May 2020-January 2021

#### Global social distancing assessment

In crude analyses, participants who reported that they *always or sometimes* engaged in social distancing with people they know had a statistically significantly lower seroincidence (IRR_always vs never_=0.30, 95% CI 0.15-0.72; IRR_sometimes vs never_=0.35, 95% CI 0.16-0.86) compared with those who said they *never* social distanced with people they know (Table 4). In multivariable analyses that controlled for sociodemographics and comorbidities (Model 1, aIRR_always vs never_=0.37, 95% CI: 0.18-0.89), and additionally for community-level transmission (Model 2, aIRR_always vs never_=0.43, 95% CI: 0.21-1.04), participants who reported *always* social distancing with those they know (versus never) had a significantly lower seroincidence, although the 95% confidence intervals were wider. Participants reporting that they socially distanced *sometimes* with people they knew also had lower incidence of seroconversion compared with those never socially distancing in both models (Table 4).

**Table 4:** Crude and adjusted* incidence rate ratios (IRRs) for seroincidence in the CHASING COVID Cohort Study, May 2020-January 2021

|  | Crude |  | Adjusted<br>(Model 1 **) |  | Adjusted<br>(Model 2 ***) |  |
| --- | --- | --- | --- | --- | --- | --- |
|  | IRR | 95% CI | IRR | 95% CI | IRR | 95% CI |
| **Social distancing with people you know (ref:Never)** |  |  |  |  |  |  |
| Always | 0.30 | 0.15, 0.72 | 0.37 | 0.18, 0.89 | 0.43 | 0.21, 1.04 |
| Sometimes | 0.35 | 0.16, 0.86 | 0.42 | 0.19, 1.05 | 0.47 | 0.22, 1.19 |
| Not applicable (as per participant) | 0.54 | 0.14, 1.80 | 0.70 | 0.18, 2.36 | 0.73 | 0.19, 2.48 |
| **Social distancing with people you do not know (ref:Never)** |  |  |  |  |  |  |
| Always | 0.54 | 0.34, 0.90 | 0.60 | 0.37, 1.00 | 0.64 | 0.39, 1.07 |
| Sometimes | 0.53 | 0.34, 0.85 | 0.57 | 0.37, 0.92 | 0.60 | 0.38, 0.97 |
| Not applicable (as per participant) | 0.30 | 0.09, 0.78 | 0.34 | 0.10, 0.89 | 0.36 | 0.10, 0.95 |
| **Composite measure of risk factors (ref:Low)** |  |  |  |  |  |  |
| Medium | 1.54 | 0.97, 2.51 | 1.61 | 1.00, 2.64 | 1.59 | 0.99, 2.60 |
| High | 3.18 | 2.11, 4.95 | 3.17 | 2.07, 5.00 | 3.12 | 2.04, 4.93 |

\* Adjusted for test sensitivity and specificity
\*\* Model 1: Adjusted for age, gender, race/ethnicity, and comorbidities
\*\*\* Model 2: Adjusted additionally for county-level changes in community level transmission

Participants who reported social distancing *always* or *sometimes (vs. never)* with people they don’t know had lower seroincidence rates in both crude analyses and adjusted models, however the association was weaker than it was for social distancing with people they *did* know, with IRRs and aIRRs ranging from 0.53-0.64.

### Clinical and public health outcomes among persons with SARS-CoV-2 seroconversion during May 2020-January 2021

Among the 161 individuals who seroconverted during May 2020-January 2021, only 27 (16.8%) were aware that they had a prior SARS-CoV-2 infection (Table 5). A substantial proportion (28.0%) recalled no symptoms of COVID-like illness (i.e., were asymptomatic cases). In terms of public health outcomes, 60.3% said that they were ever tested for SARS-CoV-2 outside the study (Table 5), but only half of them (26.7% of total) reported ever having a positive SARS-CoV-2 test. Only 29.2% said that they had *ever* isolated themselves from people outside their household because of their infection, and, among those who did not live alone, even fewer (17.4% overall) said they ever isolated themselves from others within their household. In terms of contact tracing, only 19.3% of all seroconverters were asked about contacts following diagnosis and only 11.8% had been informed by a contact tracer that they may have had contact with someone confirmed to have SARS-CoV-2. Only 5.0% of all seroconverters were told by a contact tracer to stay home for a period of time because they had COVID-19. Findings were similar among all 323 seropositive participants (data not shown).

**Table 5.** Clinical and public health outcomes among persons with seroincident SARS-CoV-2, May 2020-January 2021

|  | N | % |
| --- | --- | --- |
| **Total** | 161 | 50% |
| **Symptoms and clinical outcomes** |  |  |
| PCR confirmed diagnosis | 43 | 26.7 |
| Asymptomatic * | 45 | 28 |
| Mild (symptomatic, but didn't seek care) | 99 | 61.5 |
| Ever had COVID like illness* | 114 | 70.8 |
| Nasal discharge, congestion or sneezing | 100 | 62.1 |
| Cough/Cough up phlegm | 75 | 46.6 |
| Cough up blood | 0 | 0 |
| Sore throat | 65 | 40.4 |
| Itchy eye or eye pain | 53 | 32.9 |
| Shortness of breath or chest pain | 32 | 19.9 |
| Stomachache, diarrhea, nausea or vomiting | 67 | 41.6 |
| Rash | 12 | 7.5 |
| Loss of smell | 31 | 19.3 |
| Headache | 89 | 55.3 |
| Fever, chills or repeated chills | 44 | 27.3 |
| Myalgia | 58 | 36 |
| Ever hospitalized | 4 | 2.5 |
| **Public health outcomes and testing history** |  |  |
| Ever tested for COVID | 97 | 60.3 |
| Positive SARS-CoV-2 PCR test | 43 | 26.7 |
| Isolated from people outside household | 47 | 29.2 |
| Isolated from people within household ** | 28 | 17.4 |
| Quarantined after contact with COVID | 31 | 19.3 |
| Asked about contacts after COVID diagnosis | 31 | 19.3 |
| Told about contacts with COVID case | 19 | 11.8 |
| Encouraged to get tested because of exposure to case | 10 | 6.2 |
| Told to stay home for a period of time | 8 | 5 |

\* Based on Council of State and Territorial Epidemiologists case definition
\*\* Among those with others in the household

## DISCUSSION

We report findings from a large community-based prospective epidemiologic study of SARS-CoV-2 incidence and risk factors in the U.S. during May 2020-January 2021. Using serologic tests, we longitudinally characterized the incidence of SARS-CoV-2 infection in relation to a range of modifiable risk factors. We found that social distancing with those whom participants knew as well as those whom they did not know was highly protective against SARS-CoV-2 infection, even after controlling for other risk factors and measured confounders. Finally, public health strategies such as quarantining, testing, isolation, and contact tracing appear to have had inadequate coverage and adoption during the infectious periods of those infected with SARS-CoV-2, limiting their effectiveness at reducing SARS-CoV-2 transmission in the community. Taken together, our study findings document some of the principal reasons why the U.S. has continued to experience sustained community transmission, hospitalizations, and deaths from COVID-19 in many areas, including into the vaccine and Delta variant eras.

The protective association observed for two global measures of social distancing persisted even after controlling for other differences. This protective association was strongest for participants reporting that they always or sometimes socially distanced from people whom they knew, but was also present among those who always or sometimes socially distanced from those they did not know. Our findings suggest a need for more effective and consistent messaging around social distancing.

We observed substantially increased risk, in dose response fashion, from a number of other key epidemiologic risk factors and exposures reflected in a composite risk score. Among participants in the top tertile of the risk score the risk of seroincidence was 3-fold higher, accounting for 55% of the observed seroconversions. Reducing multiple risk factors (e.g., through policies on masking, mass gatherings, indoor dining/bars, social distancing, air travel) would likely substantially reduce community transmission.

Our findings suggest that elevated risk among essential workers, observed early in the U.S. pandemic, persisted into the second phase of the pandemic. Essential workers risk exposure to SARS-CoV-2 not only in their workplaces, but also in their communities and as part of their commutes to and from work, especially when using public transportation. The increased burden of risk of SARS-CoV-2 infection in essential workers is shared with their household members, among whom transmission is very efficient.[28] National, state and local workplace safety measures, mandates, and policies that protect essential workers have the added benefit of protecting household members and other close contacts of workers.

Finally, detailed examination of 161 individuals with SARS-CoV-2 seroconversion showed major gaps in the reach of public health interventions aimed at reducing the spread of SARS-CoV-2. Most who seroconverted (73.3%) did not report a prior positive PCR test, and a substantial proportion were asymptomatic. Moreover, few people who seroconverted in our study reported being reached by contact tracers (11.8%). These results highlight the barriers to successful implementation of isolation, contact tracing, and quarantine. Now that rapid home tests are easily available, frequent proactive testing at home can be a more effective way to capture asymptomatic and presymptomatic cases early and prevent onward transmission.

Our study, as well as data from routine case surveillance, highlight that the drivers of racial/ethnic disparities in SARS-CoV-2 risk have not been addressed in the U.S. The ultimate drivers of these disparities need to be targeted by governments, health departments and researchers, and used to course-correct the public health response.[29] Structural factors, such as household crowding, the need to go to work to avoid income loss, and inequitable access to SARS-CoV-2 testing[30], create and perpetuate a disparate burden of SARS-CoV-2 exposure and incidence.[31] To date, no targeted strategies or policies have been deployed that aim to protect those who cannot afford missing work, including, but not only, essential workers. Public health leaders and policy makers should anticipate and proactively design pandemic response implementation strategies, with performance metrics related to inequalities[32,33], that account for and counteract the prevailing structural forces, including structural racism, that create, maintain, or exacerbate inequities in safety, health, and well-being during and after a public health crisis.[32–35]

Our study has limitations worth noting. The observed cumulative incidence in our cohort may be lower than the true cumulative incidence in our cohort because of waning of SARS-CoV-2 antibodies.[36] Recent studies suggest waning of antibodies to both nucleocapsid and spike proteins.[5] Combined with the timing of specimen collection relative to infection for many participants in our cohort (median of 190 days)[12], this could mean that we have underestimated the true cumulative incidence. Next, estimated associations between SARS-CoV-2 risk factors and incidence are subject to confounding. The crude associations we presented may also vary by setting, with interpretation for some associations further hampered by small sample sizes. Some risk behaviors may have been underreported, due to social desirability, which would bias observed associations toward the null. Finally, our study period for the current analysis pre-dated the vaccine era and the emergence of the highly transmissible and possibly more virulent SARS-CoV-2 Delta variant. We could not, therefore, examine risk factors for infection among vaccinated persons. However, given recent data showing decreasing vaccine effectiveness against the Delta variant for the outcome of infection[37,38], as well as major outbreaks and high viral loads among fully vaccinated persons[3,39], it is likely that many of our findings related to transmission risk factors also apply to vaccinated persons, as they remain at risk for breakthrough infection and onward transmission of SARS-CoV-2 when engaging in some of the same risk factors. A similar study performed in the Delta variant era might result in stronger risk factor and weaker protective factor associations with seroconversion, as well as a different estimate of proportion asymptomatic.

## Conclusion

Modifiable risk factors and poor reach of public health strategies continue to drive transmission of SARS-CoV-2 across the U.S. While continuing to increase vaccine access and coverage, it remains critical for public health agencies to simultaneously reduce risk factors and address structural factors that contribute to high incidence and persistent inequities. Future research will include monitoring SARS-CoV-2 outcomes, including infections in the vaccine and Delta variant eras, at least through December 2021.

## Data Availability

The data that support the findings of this study are available on request from the corresponding author, [DN]. The data are not yet publicly available, but we are preparing to post a deidentified, HIPAA compliant, public use version of our baseline and follow-up data on GitHub.

https://cunyisph.org/chasing-covid/

## CONTRIBUTORS

DN, MSR, SGK, MMR, conceptualized the study. MSR, MC and DN performed statistical analyses. DN and MSR wrote the first draft of the paper. DN, MSR, MC, SGK, and AP contributed to interpreting the data, DN, RZ, MSR, MC, SGK, WY, AB, CM, SK, AM, MMR, DAW, AP, LW, and CG contributed to the writing and revising of the manuscript. SGK, WY, AB, CM, SK, and DN contributed to data collection, cleaning and management. DN, SGK, MMR and CG contributed to obtaining funding for the research.

## ACKNOWLEDGEMENTS

The authors wish to thank the participants of the CHASING COVID Cohort Study. We are grateful to you for your contributions to the advancement of science around the SARS-CoV-2 pandemic. We thank Prof. Patrick Sullivan and MTL for local validation work on the serologic assays for use with DBS that greatly benefited our study. We are also grateful to MTL Labs for processing specimen collection kits and serologic testing of our cohort’s specimens.

## FUNDING

Funding for this project is provided by The National Institute of Allergy and Infectious Diseases (NIAID), award number 3UH3AI133675-04S1 (MPIs: D Nash and C Grov), the CUNY Institute for Implementation Science in Population Health (cunyisph.org) and the COVID-19 Grant Program of the CUNY Graduate School of Public Health and Health Policy, and National Institute of Child Health and Human Development grant P2C HD050924 (Carolina Population Center).

## STATISTICAL APPENDIX

### LASSO

The study sample of 3,422 participants with 2 serosamples was split randomly into equally sized training and test data sets. A grouped LASSO regression was fit using training data and 10-fold cross validation was used to obtain the minimum value of lambda (the tuning parameter). The variables included in the LASSO model were household crowding, having children in the household, confirmed COVID-19 case in a household member, attending mass gatherings, indoor dining in a restaurant/bar, outdoor dining in a restaurant/bar, visiting places of worship, visiting public park or pool, gathering with groups of 10 or more indoors and/or outdoors, mask use while grocery shopping, mask use while visiting non-household members, mask use indoors in gym/salons, mask use indoors at work, mask use outdoors, using public transit, recent air travel, alcohol use,and substance use. Using the minimum lambda value, a grouped LASSO model was run on the entire dataset (training + test) to obtain factors that best predicted seroconversion. The LASSO model selected having a confirmed COVID-19 case in a household member, indoor dining in a bar/restaurant, gathering with groups of 10 or more outdoors, and mask use indoors in salons/gyms as the most predictive of seroconversion in our cohort. A logistic regression model was fitted using these variables selected by LASSO regression with seroconversion (Y/N) as the outcome. Coefficients of the variables were multiplied by 10 and rounded up to the nearest integer to create a score associated with that variable/risk factor. If a participant engaged in a given risk factor, they were assigned the score associated with that risk factor. Scores were then summed across risk factors, normalized between 0 and 100, and a total composite risk score was created for each participant.

### County-level COVID-19 community transmission

We used population-based, county-level death rates, lagged by 23 days [22,23], as a proxy for community transmission. We assumed that data on the number of deaths for a given day represented community transmission that was occurring 23 days earlier, specifically 5 days from infection to symptom onset (reflecting the average incubation period) [24]; 5 days from symptom onset to severe disease/hospitalization [25]; and 13 days from hospitalization to death. [26]

We obtained the cumulative number of COVID-19 deaths per 100,000 persons for each county, using data from the New York Times Github website (from 01/21/2020 to 01/07/2021)[27] on the days samples were collected in Period 1 and Period 2 for each participant. We calculated the rate of change in cumulative COVID-19 deaths per 100,000 at three time points for each participant, including between: 1) January 2020 (approximate start of U.S. pandemic) and the date of specimen collection in Period 1; 2) the start of the pandemic and the date of specimen collection in Period 2; and 3) the difference between the specimen collection dates in Period 1 and Period 2. The rate of change in COVID-19 deaths per 100,000 were divided into tertiles of low, medium and high community transmission.

## Supplemental Material

**Table S1.** Baseline characteristics and serologic testing of CHASING COVID Cohort Study participants by time period of testing

|  | All participants |  | Participants with one or more serologic test (May 2020-January 2021) |  | Participants with a serologic test in Period 1 (May-September 2020) |  | Participants with a serologic test in Period 2 (November 2020-January 2021) |  |
| --- | --- | --- | --- | --- | --- | --- | --- | --- |
|  | N | % | N | % | N | % | N | % |
| **Total** | 6,738 | 100% | 4510 | 67% | 4232 | 63% | 3883 | 58% |
| **Age** |  |  |  |  |  |  |  |  |
| Median (IQR) | 37 (29,51) |  | 41 (31, 55) |  | 41, (32, 56) |  | 41, (32, 56) |  |
| 18-29 | 1802 | 26.74 | 876 | 19.4 | 800 | 18.9 | 722 | 18.6 |
| 30-39 | 1955 | 29.01 | 1253 | 27.8 | 1171 | 27.7 | 1074 | 27.7 |
| 40-49 | 1163 | 17.26 | 858 | 19.0 | 812 | 19.2 | 738 | 19.0 |
| 50-59 | 792 | 11.75 | 658 | 14.6 | 620 | 14.7 | 582 | 15.0 |
| 60+ | 1026 | 15.23 | 865 | 19.2 | 829 | 19.6 | 767 | 19.8 |
| **Gender** |  |  |  |  |  |  |  |  |
| Male | 3042 | 45.15 | 2018 | 44.8 | 1898 | 44.9 | 1738 | 44.8 |
| Female | 3525 | 52.32 | 2360 | 52.3 | 2211 | 52.2 | 2038 | 52.5 |
| Non-Binary/Transgender | 171 | 2.54 | 132 | 2.9 | 123 | 2.9 | 107 | 2.8 |
| **Race/Ethnicity** |  |  |  |  |  |  |  |  |
| Non-Hispanic White | 3829 | 56.83 | 2966 | 65.8 | 2818 | 66.6 | 2568 | 66.1 |
| Hispanic | 1308 | 19.41 | 679 | 15.1 | 622 | 14.7 | 584 | 15.0 |
| Non-Hispanic Black | 848 | 12.59 | 378 | 8.4 | 341 | 8.1 | 316 | 8.1 |
| Asian | 439 | 6.52 | 286 | 6.3 | 262 | 6.2 | 245 | 6.3 |
| Pacific Islander | 36 | 0.53 | 18 | 0.4 | 18 | 0.4 | 16 | 0.4 |
| Other | 250 | 3.71 | 168 | 3.7 | 157 | 3.7 | 141 | 3.6 |
| Missing | 28 | 0.42 | 15 | 0.3 | 14 | 0.3 | 13 | 0.3 |
| **Education** |  |  |  |  |  |  |  |  |
| Less than high school | 123 | 1.83 | 54 | 1.2 | 47 | 1.1 | 48 | 1.2 |
| High school graduate | 875 | 12.99 | 403 | 8.9 | 365 | 8.6 | 331 | 8.5 |
| Some college | 1888 | 28.02 | 1151 | 25.5 | 1052 | 24.9 | 954 | 24.6 |
| College graduate | 3852 | 57.17 | 2902 | 64.4 | 2768 | 65.4 | 2550 | 65.7 |
| **Employment** |  |  |  |  |  |  |  |  |
| Employed | 4246 | 63.02 | 2774 | 61.5 | 2612 | 61.7 | 2394 | 61.7 |
| Out of work | 830 | 12.32 | 544 | 12.1 | 503 | 11.9 | 461 | 11.9 |
| Homemaker | 352 | 5.22 | 242 | 5.4 | 225 | 5.3 | 203 | 5.2 |
| Student | 608 | 9.02 | 367 | 8.1 | 329 | 7.8 | 305 | 7.9 |
| Retired | 702 | 10.42 | 583 | 12.9 | 563 | 13.3 | 520 | 13.4 |
| **Household income** |  |  |  |  |  |  |  |  |
| Less than \$35,000 | 1967 | 29.19 | 1186 | 26.3 | 1112 | 26.3 | 987 | 25.4 |
| \$35-49,999 | 753 | 11.18 | 496 | 11.0 | 457 | 10.8 | 440 | 11.3 |
| \$50-69,999 | 959 | 14.23 | 643 | 14.3 | 600 | 14.2 | 568 | 14.6 |
| \$70-99,999 | 1058 | 15.7 | 741 | 16.4 | 706 | 16.7 | 660 | 17.0 |
| \$100,000+ | 1793 | 26.61 | 1329 | 29.5 | 1258 | 29.7 | 1126 | 29.0 |
| Missing | 208 | 3.08 | 115 | 2.6 | 99 | 2.3 | 102 | 2.6 |
| **Setting** |  |  |  |  |  |  |  |  |
| Urban | 2835 | 42.07 | 1911 | 42.4 | 1800 | 42.5 | 1642 | 42.3 |
| Suburban | 1792 | 26.59 | 1186 | 26.3 | 1119 | 26.4 | 1020 | 26.3 |
| Rural | 2049 | 30.41 | 1412 | 31.3 | 1312 | 31.0 | 1220 | 31.4 |
| Missing | 62 | 0.92 | 1 | 0.0 | 1 | 0.0 | 1 | 0.0 |
| **Geographic region** |  |  |  |  |  |  |  |  |
| Northeast | 1891 | 28.06 | 1320 | 29.3 | 1253 | 29.6 | 1124 | 29.0 |
| Midwest | 1130 | 16.77 | 805 | 17.9 | 763 | 18.0 | 699 | 18.0 |
| South | 2040 | 30.28 | 1269 | 28.1 | 1168 | 27.6 | 1102 | 28.4 |
| West | 1630 | 24.19 | 1111 | 24.6 | 1043 | 24.7 | 954 | 24.6 |
| US Territories | 8 | 0.12 | 5 | 0.1 | 5 | 0.1 | 4 | 0.1 |
| Missing | 39 | 0.58 | 0 | 0.0 | 0 | 0.0 | 0 | 0.0 |
| **Healthcare worker** |  |  |  |  |  |  |  |  |
| No | 5859 | 86.95 | 4048 | 89.8 | 3797 | 89.7 | 3492 | 89.9 |
| Yes | 795 | 11.8 | 425 | 9.4 | 400 | 9.5 | 355 | 9.1 |
| Missing | 84 | 1.25 | 37 | 0.8 | 35 | 0.8 | 36 | 0.9 |
| **Essential worker \*** |  |  |  |  |  |  |  |  |
| No | 4785 | 71.02 | 3170 | 70.3 | 2992 | 70.7 | 2700 | 69.5 |
| Yes | 1953 | 28.98 | 1340 | 29.7 | 1240 | 29.3 | 1183 | 30.5 |
| **Higher risk for severe COVID \*\*** |  |  |  |  |  |  |  |  |
| No | 3165 | 46.97 | 2097 | 46.5 | 1973 | 46.6 | 1797 | 46.3 |
| Yes | 3573 | 53.03 | 2413 | 53.5 | 2259 | 53.4 | 2086 | 53.7 |

\* Combined from three follow-up interviews between August and November 2020
\*\* >60 years old, or reported co-morbidity, or current smoker. Comorbidity was defined as having history of heart attack, depression, angina, immunosuppression, type 2 diabetes, high blood pressure, cancer, asthma, COPD, chronic kidney disease, and/or HIV/AIDS

**Table S2.**
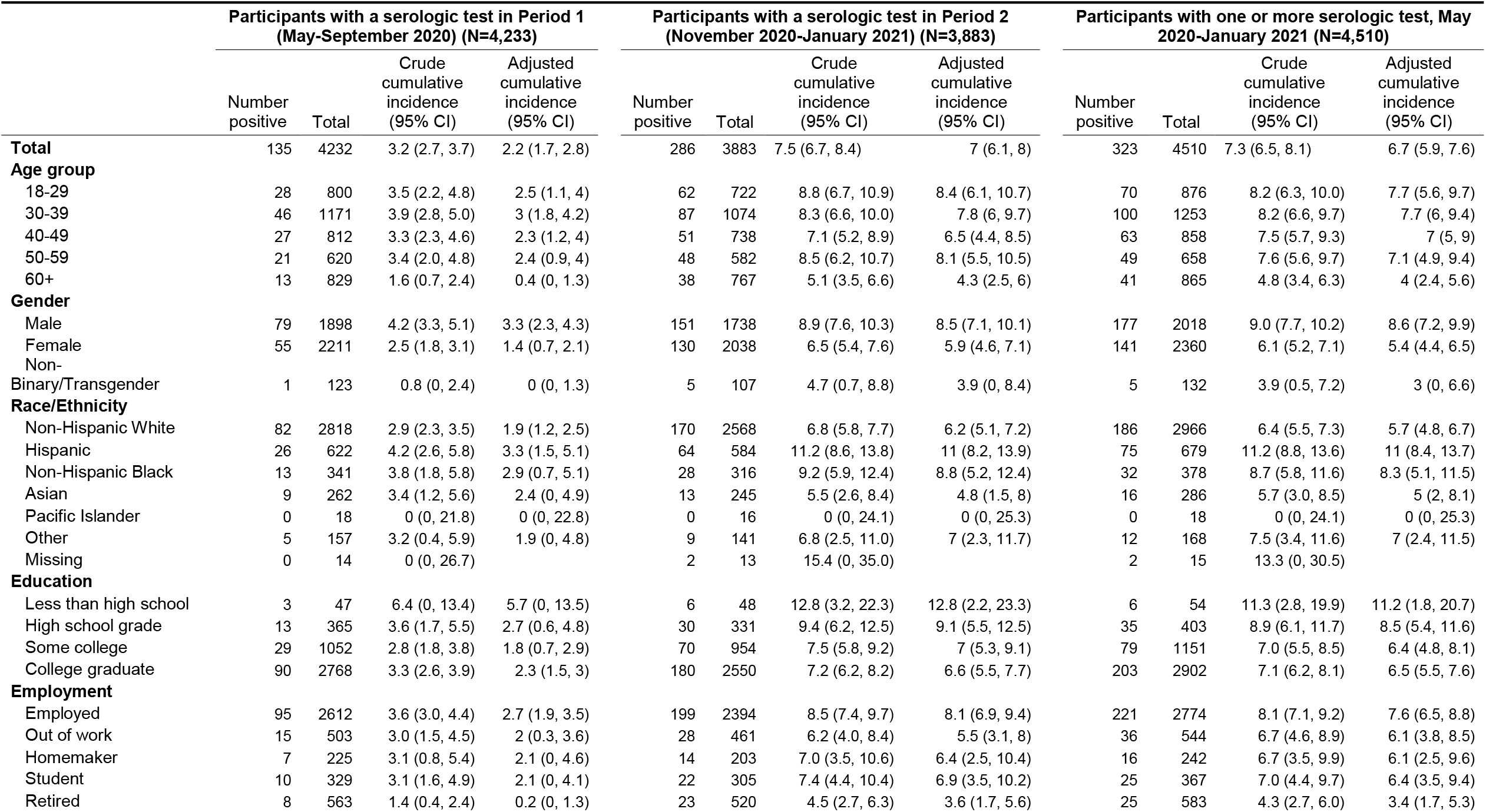
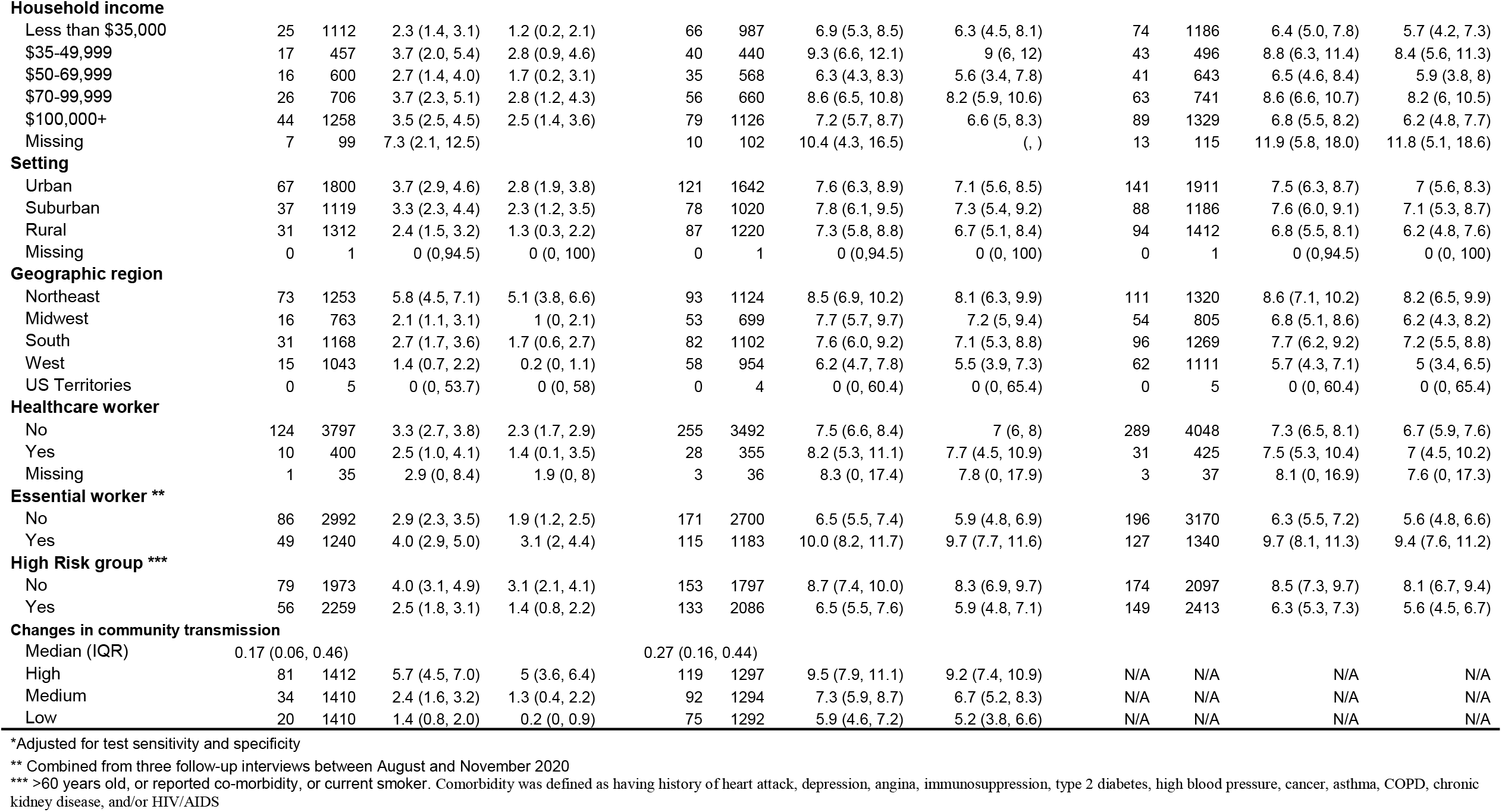
Crude and adjusted* serology-based cumulative incidence estimates of SARS-CoV-2 infection in CHASING COVID Cohort Study participants by time period of testing

|  | Participants with a serologic test in Period 1<br>(May-September 2020) (N=4,233) |  |  |  | Participants with a serologic test in Period 2<br>(November 2020-January 2021) (N=3,883) |  |  |  | Participants with one or more serologic test, May<br>2020-January 2021 (N=4,510) |  |  |  |
| --- | --- | --- | --- | --- | --- | --- | --- | --- | --- | --- | --- | --- |
|  | Number<br>positive | Total | Crude<br>cumulative<br>incidence<br>(95% CI) | Adjusted<br>cumulative<br>incidence<br>(95% CI) | Number<br>positive | Total | Crude<br>cumulative<br>incidence<br>(95% CI) | Adjusted<br>cumulative<br>incidence<br>(95% CI) | Number<br>positive | Total | Crude<br>cumulative<br>incidence<br>(95% CI) | Adjusted<br>cumulative<br>incidence<br>(95% CI) |
| **Total** | 135 | 4232 | 3.2 (2.7, 3.7) | 2.2 (1.7, 2.8) | 286 | 3883 | 7.5 (6.7, 8.4) | 7 (6.1, 8) | 323 | 4510 | 7.3 (6.5, 8.1) | 6.7 (5.9, 7.6) |
| **Age group** |  |  |  |  |  |  |  |  |  |  |  |  |
| 18-29 | 28 | 800 | 3.5 (2.2, 4.8) | 2.5 (1.1, 4) | 62 | 722 | 8.8 (6.7, 10.9) | 8.4 (6.1, 10.7) | 70 | 876 | 8.2 (6.3, 10.0) | 7.7 (5.6, 9.7) |
| 30-39 | 46 | 1171 | 3.9 (2.8, 5.0) | 3 (1.8, 4.2) | 87 | 1074 | 8.3 (6.6, 10.0) | 7.8 (6, 9.7) | 100 | 1253 | 8.2 (6.6, 9.7) | 7.7 (6, 9.4) |
| 40-49 | 27 | 812 | 3.3 (2.3, 4.6) | 2.3 (1.2, 4) | 51 | 738 | 7.1 (5.2, 8.9) | 6.5 (4.4, 8.5) | 63 | 858 | 7.5 (5.7, 9.3) | 7 (5, 9) |
| 50-59 | 21 | 620 | 3.4 (2.0, 4.8) | 2.4 (0.9, 4) | 48 | 582 | 8.5 (6.2, 10.7) | 8.1 (5.5, 10.5) | 49 | 658 | 7.6 (5.6, 9.7) | 7.1 (4.9, 9.4) |
| 60+ | 13 | 829 | 1.6 (0.7, 2.4) | 0.4 (0, 1.3) | 38 | 767 | 5.1 (3.5, 6.6) | 4.3 (2.5, 6) | 41 | 865 | 4.8 (3.4, 6.3) | 4 (2.4, 5.6) |
| **Gender** |  |  |  |  |  |  |  |  |  |  |  |  |
| Male | 79 | 1898 | 4.2 (3.3, 5.1) | 3.3 (2.3, 4.3) | 151 | 1738 | 8.9 (7.6, 10.3) | 8.5 (7.1, 10.1) | 177 | 2018 | 9.0 (7.7, 10.2) | 8.6 (7.2, 9.9) |
| Female | 55 | 2211 | 2.5 (1.8, 3.1) | 1.4 (0.7, 2.1) | 130 | 2038 | 6.5 (5.4, 7.6) | 5.9 (4.6, 7.1) | 141 | 2360 | 6.1 (5.2, 7.1) | 5.4 (4.4, 6.5) |
| Non-Binary/Transgender | 1 | 123 | 0.8 (0, 2.4) | 0 (0, 1.3) | 5 | 107 | 4.7 (0.7, 8.8) | 3.9 (0, 8.4) | 5 | 132 | 3.9 (0.5, 7.2) | 3 (0, 6.6) |
| **Race/Ethnicity** |  |  |  |  |  |  |  |  |  |  |  |  |
| Non-Hispanic White | 82 | 2818 | 2.9 (2.3, 3.5) | 1.9 (1.2, 2.5) | 170 | 2568 | 6.8 (5.8, 7.7) | 6.2 (5.1, 7.2) | 186 | 2966 | 6.4 (5.5, 7.3) | 5.7 (4.8, 6.7) |
| Hispanic | 26 | 622 | 4.2 (2.6, 5.8) | 3.3 (1.5, 5.1) | 64 | 584 | 11.2 (8.6, 13.8) | 11 (8.2, 13.9) | 75 | 679 | 11.2 (8.8, 13.6) | 11 (8.4, 13.7) |
| Non-Hispanic Black | 13 | 341 | 3.8 (1.8, 5.8) | 2.9 (0.7, 5.1) | 28 | 316 | 9.2 (5.9, 12.4) | 8.8 (5.2, 12.4) | 32 | 378 | 8.7 (5.8, 11.6) | 8.3 (5.1, 11.5) |
| Asian | 9 | 262 | 3.4 (1.2, 5.6) | 2.4 (0, 4.9) | 13 | 245 | 5.5 (2.6, 8.4) | 4.8 (1.5, 8) | 16 | 286 | 5.7 (3.0, 8.5) | 5 (2, 8.1) |
| Pacific Islander | 0 | 18 | 0 (0, 21.8) | 0 (0, 22.8) | 0 | 16 | 0 (0, 24.1) | 0 (0, 25.3) | 0 | 18 | 0 (0, 24.1) | 0 (0, 25.3) |
| Other | 5 | 157 | 3.2 (0.4, 5.9) | 1.9 (0, 4.8) | 9 | 141 | 6.8 (2.5, 11.0) | 7 (2.3, 11.7) | 12 | 168 | 7.5 (3.4, 11.6) | 7 (2.4, 11.5) |
| Missing | 0 | 14 | 0 (0, 26.7) |  | 2 | 13 | 15.4 (0, 35.0) |  | 2 | 15 | 13.3 (0, 30.5) |  |
| **Education** |  |  |  |  |  |  |  |  |  |  |  |  |
| Less than high school | 3 | 47 | 6.4 (0, 13.4) | 5.7 (0, 13.5) | 6 | 48 | 12.8 (3.2, 22.3) | 12.8 (2.2, 23.3) | 6 | 54 | 11.3 (2.8, 19.9) | 11.2 (1.8, 20.7) |
| High school grade | 13 | 365 | 3.6 (1.7, 5.5) | 2.7 (0.6, 4.8) | 30 | 331 | 9.4 (6.2, 12.5) | 9.1 (5.5, 12.5) | 35 | 403 | 8.9 (6.1, 11.7) | 8.5 (5.4, 11.6) |
| Some college | 29 | 1052 | 2.8 (1.8, 3.8) | 1.8 (0.7, 2.9) | 70 | 954 | 7.5 (5.8, 9.2) | 7 (5.3, 9.1) | 79 | 1151 | 7.0 (5.5, 8.5) | 6.4 (4.8, 8.1) |
| College graduate | 90 | 2768 | 3.3 (2.6, 3.9) | 2.3 (1.5, 3) | 180 | 2550 | 7.2 (6.2, 8.2) | 6.6 (5.5, 7.7) | 203 | 2902 | 7.1 (6.2, 8.1) | 6.5 (5.5, 7.6) |
| **Employment** |  |  |  |  |  |  |  |  |  |  |  |  |
| Employed | 95 | 2612 | 3.6 (3.0, 4.4) | 2.7 (1.9, 3.5) | 199 | 2394 | 8.5 (7.4, 9.7) | 8.1 (6.9, 9.4) | 221 | 2774 | 8.1 (7.1, 9.2) | 7.6 (6.5, 8.8) |
| Out of work | 15 | 503 | 3.0 (1.5, 4.5) | 2 (0.3, 3.6) | 28 | 461 | 6.2 (4.0, 8.4) | 5.5 (3.1, 8) | 36 | 544 | 6.7 (4.6, 8.9) | 6.1 (3.8, 8.5) |
| Homemaker | 7 | 225 | 3.1 (0.8, 5.4) | 2.1 (0, 4.6) | 14 | 203 | 7.0 (3.5, 10.6) | 6.4 (2.5, 10.4) | 16 | 242 | 6.7 (3.5, 9.9) | 6.1 (2.5, 9.6) |
| Student | 10 | 329 | 3.1 (1.6, 4.9) | 2.1 (0, 4.1) | 22 | 305 | 7.4 (4.4, 10.4) | 6.9 (3.5, 10.2) | 25 | 367 | 7.0 (4.4, 9.7) | 6.4 (3.5, 9.4) |
| Retired | 8 | 563 | 1.4 (0.4, 2.4) | 0.2 (0, 1.3) | 23 | 520 | 4.5 (2.7, 6.3) | 3.6 (1.7, 5.6) | 25 | 583 | 4.3 (2.7, 6.0) | 3.4 (1.7, 5.3) |

|  |  |  |  |  |  |  |  |  |  |  |  |  |  |
| --- | --- | --- | --- | --- | --- | --- | --- | --- | --- | --- | --- | --- | --- |
| Household income |  |  |  |  |  |  |  |  |  |  |  |  |  |
| Less than \$35,000 | 25 | 1112 | 2.3 (1.4, 3.1) | 1.2 (0.2, 2.1) | 66 | 987 | 6.9 (5.3, 8.5) | 6.3 (4.5, 8.1) | 74 | 1186 | 6.4 (5.0, 7.8) | 5.7 (4.2, 7.3) | |
| \$35-49,999 | 17 | 457 | 3.7 (2.0, 5.4) | 2.8 (0.9, 4.6) | 40 | 440 | 9.3 (6.6, 12.1) | 9 (6, 12) | 43 | 496 | 8.8 (6.3, 11.4) | 8.4 (5.6, 11.3) | |
| \$50-69,999 | 16 | 600 | 2.7 (1.4, 4.0) | 1.7 (0.2, 3.1) | 35 | 568 | 6.3 (4.3, 8.3) | 5.6 (3.4, 7.8) | 41 | 643 | 6.5 (4.6, 8.4) | 5.9 (3.8, 8) | |
| \$70-99,999 | 26 | 706 | 3.7 (2.3, 5.1) | 2.8 (1.2, 4.3) | 56 | 660 | 8.6 (6.5, 10.8) | 8.2 (5.9, 10.6) | 63 | 741 | 8.6 (6.6, 10.7) | 8.2 (6, 10.5) | |
| \$100,000+ | 44 | 1258 | 3.5 (2.5, 4.5) | 2.5 (1.4, 3.6) | 79 | 1126 | 7.2 (5.7, 8.7) | 6.6 (5, 8.3) | 89 | 1329 | 6.8 (5.5, 8.2) | 6.2 (4.8, 7.7) | |
| Missing | 7 | 99 | 7.3 (2.1, 12.5) |  | 10 | 102 | 10.4 (4.3, 16.5) | (, ) | 13 | 115 | 11.9 (5.8, 18.0) | 11.8 (5.1, 18.6) |  |
| Setting |  |  |  |  |  |  |  |  |  |  |  |  |  |
| Urban | 67 | 1800 | 3.7 (2.9, 4.6) | 2.8 (1.9, 3.8) | 121 | 1642 | 7.6 (6.3, 8.9) | 7.1 (5.6, 8.5) | 141 | 1911 | 7.5 (6.3, 8.7) | 7 (5.6, 8.3) |  |
| Suburban | 37 | 1119 | 3.3 (2.3, 4.4) | 2.3 (1.2, 3.5) | 78 | 1020 | 7.8 (6.1, 9.5) | 7.3 (5.4, 9.2) | 88 | 1186 | 7.6 (6.0, 9.1) | 7.1 (5.3, 8.7) |  |
| Rural | 31 | 1312 | 2.4 (1.5, 3.2) | 1.3 (0.3, 2.2) | 87 | 1220 | 7.3 (5.8, 8.8) | 6.7 (5.1, 8.4) | 94 | 1412 | 6.8 (5.5, 8.1) | 6.2 (4.8, 7.6) |  |
| Missing | 0 | 1 | 0 (0,94.5) | 0 (0, 100) | 0 | 1 | 0 (0,94.5) | 0 (0, 100) | 0 | 1 | 0 (0,94.5) | 0 (0, 100) |  |
| Geographic region |  |  |  |  |  |  |  |  |  |  |  |  |  |
| Northeast | 73 | 1253 | 5.8 (4.5, 7.1) | 5.1 (3.8, 6.6) | 93 | 1124 | 8.5 (6.9, 10.2) | 8.1 (6.3, 9.9) | 111 | 1320 | 8.6 (7.1, 10.2) | 8.2 (6.5, 9.9) |  |
| Midwest | 16 | 763 | 2.1 (1.1, 3.1) | 1 (0, 2.1) | 53 | 699 | 7.7 (5.7, 9.7) | 7.2 (5, 9.4) | 54 | 805 | 6.8 (5.1, 8.6) | 6.2 (4.3, 8.2) |  |
| South | 31 | 1168 | 2.7 (1.7, 3.6) | 1.7 (0.6, 2.7) | 82 | 1102 | 7.6 (6.0, 9.2) | 7.1 (5.3, 8.8) | 96 | 1269 | 7.7 (6.2, 9.2) | 7.2 (5.5, 8.8) |  |
| West | 15 | 1043 | 1.4 (0.7, 2.2) | 0.2 (0, 1.1) | 58 | 954 | 6.2 (4.7, 7.8) | 5.5 (3.9, 7.3) | 62 | 1111 | 5.7 (4.3, 7.1) | 5 (3.4, 6.5) |  |
| US Territories | 0 | 5 | 0 (0, 53.7) | 0 (0, 58) | 0 | 4 | 0 (0, 60.4) | 0 (0, 65.4) | 0 | 5 | 0 (0, 60.4) | 0 (0, 65.4) |  |
| Healthcare worker |  |  |  |  |  |  |  |  |  |  |  |  |  |
| No | 124 | 3797 | 3.3 (2.7, 3.8) | 2.3 (1.7, 2.9) | 255 | 3492 | 7.5 (6.6, 8.4) | 7 (6, 8) | 289 | 4048 | 7.3 (6.5, 8.1) | 6.7 (5.9, 7.6) |  |
| Yes | 10 | 400 | 2.5 (1.0, 4.1) | 1.4 (0.1, 3.5) | 28 | 355 | 8.2 (5.3, 11.1) | 7.7 (4.5, 10.9) | 31 | 425 | 7.5 (5.3, 10.4) | 7 (4.5, 10.2) |  |
| Missing | 1 | 35 | 2.9 (0, 8.4) | 1.9 (0, 8) | 3 | 36 | 8.3 (0, 17.4) | 7.8 (0, 17.9) | 3 | 37 | 8.1 (0, 16.9) | 7.6 (0, 17.3) |  |
| Essential worker ** |  |  |  |  |  |  |  |  |  |  |  |  |  |
| No | 86 | 2992 | 2.9 (2.3, 3.5) | 1.9 (1.2, 2.5) | 171 | 2700 | 6.5 (5.5, 7.4) | 5.9 (4.8, 6.9) | 196 | 3170 | 6.3 (5.5, 7.2) | 5.6 (4.8, 6.6) |  |
| Yes | 49 | 1240 | 4.0 (2.9, 5.0) | 3.1 (2, 4.4) | 115 | 1183 | 10.0 (8.2, 11.7) | 9.7 (7.7, 11.6) | 127 | 1340 | 9.7 (8.1, 11.3) | 9.4 (7.6, 11.2) |  |
| High Risk group *** |  |  |  |  |  |  |  |  |  |  |  |  |  |
| No | 79 | 1973 | 4.0 (3.1, 4.9) | 3.1 (2.1, 4.1) | 153 | 1797 | 8.7 (7.4, 10.0) | 8.3 (6.9, 9.7) | 174 | 2097 | 8.5 (7.3, 9.7) | 8.1 (6.7, 9.4) |  |
| Yes | 56 | 2259 | 2.5 (1.8, 3.1) | 1.4 (0.8, 2.2) | 133 | 2086 | 6.5 (5.5, 7.6) | 5.9 (4.8, 7.1) | 149 | 2413 | 6.3 (5.3, 7.3) | 5.6 (4.5, 6.7) |  |
| Changes in community transmission |  |  |  |  |  |  |  |  |  |  |  |  |  |
| Median (IQR) | 0.17 (0.06, 0.46) |  |  |  | 0.27 (0.16, 0.44) |  |  |  |  |  |  |  |  |
| High | 81 | 1412 | 5.7 (4.5, 7.0) | 5 (3.6, 6.4) | 119 | 1297 | 9.5 (7.9, 11.1) | 9.2 (7.4, 10.9) | N/A | N/A | N/A | N/A |  |
| Medium | 34 | 1410 | 2.4 (1.6, 3.2) | 1.3 (0.4, 2.2) | 92 | 1294 | 7.3 (5.9, 8.7) | 6.7 (5.2, 8.3) | N/A | N/A | N/A | N/A |  |
| Low | 20 | 1410 | 1.4 (0.8, 2.0) | 0.2 (0, 0.9) | 75 | 1292 | 5.9 (4.6, 7.2) | 5.2 (3.8, 6.6) | N/A | N/A | N/A | N/A |  |

\*Adjusted for test sensitivity and specificity
\*\* Combined from three follow-up interviews between August and November 2020
\*\*\* >60 years old, or reported co-morbidity, or current smoker. Comorbidity was defined as having history of heart attack, depression, angina, immunosuppression, type 2 diabetes, high blood pressure, cancer, asthma, COPD, chronic kidney disease, and/or HIV/AIDS

**Figure S1.**
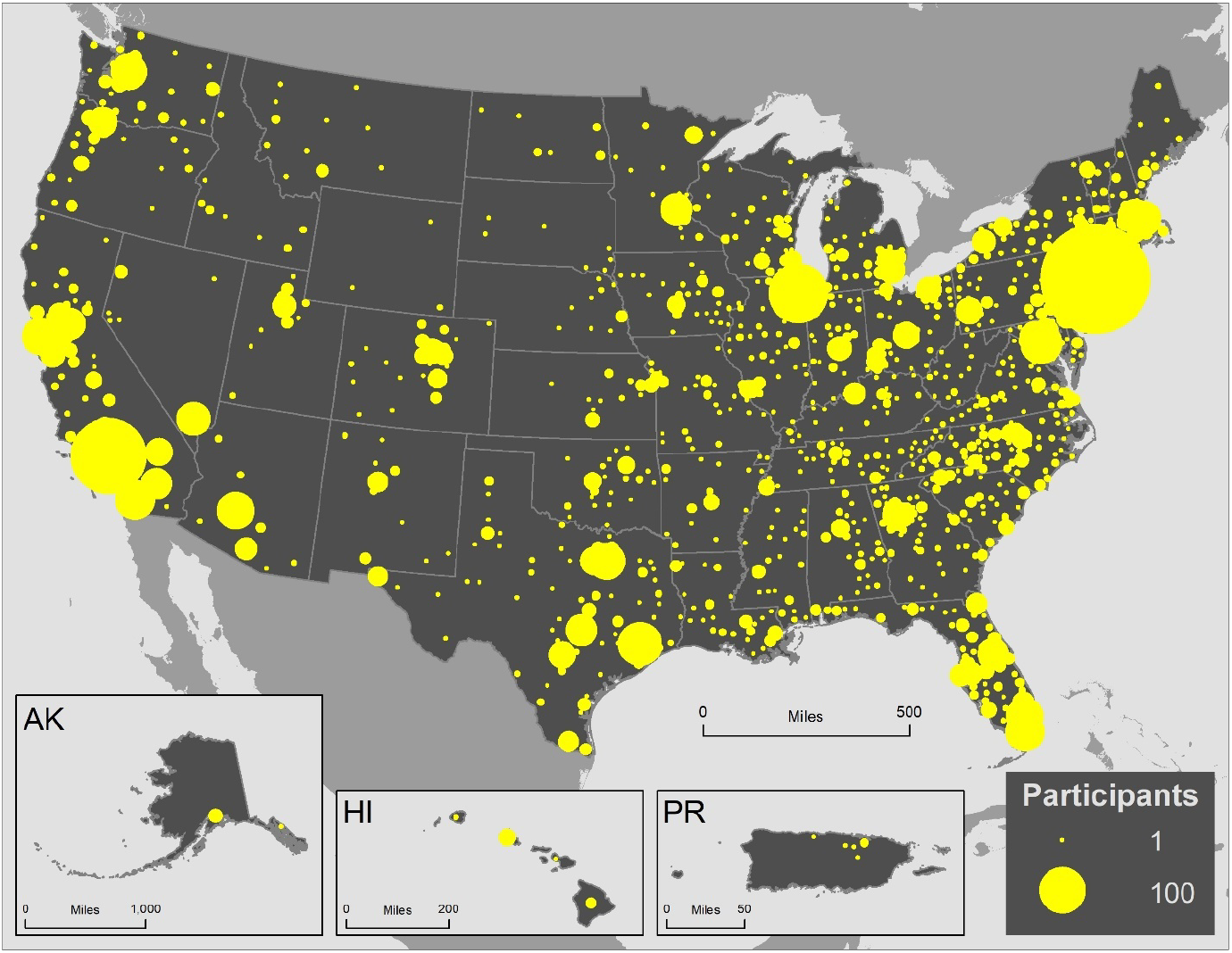
Geographic distribution of CHASING COVID Cohort participants, N=6,738

**Figure S2.**
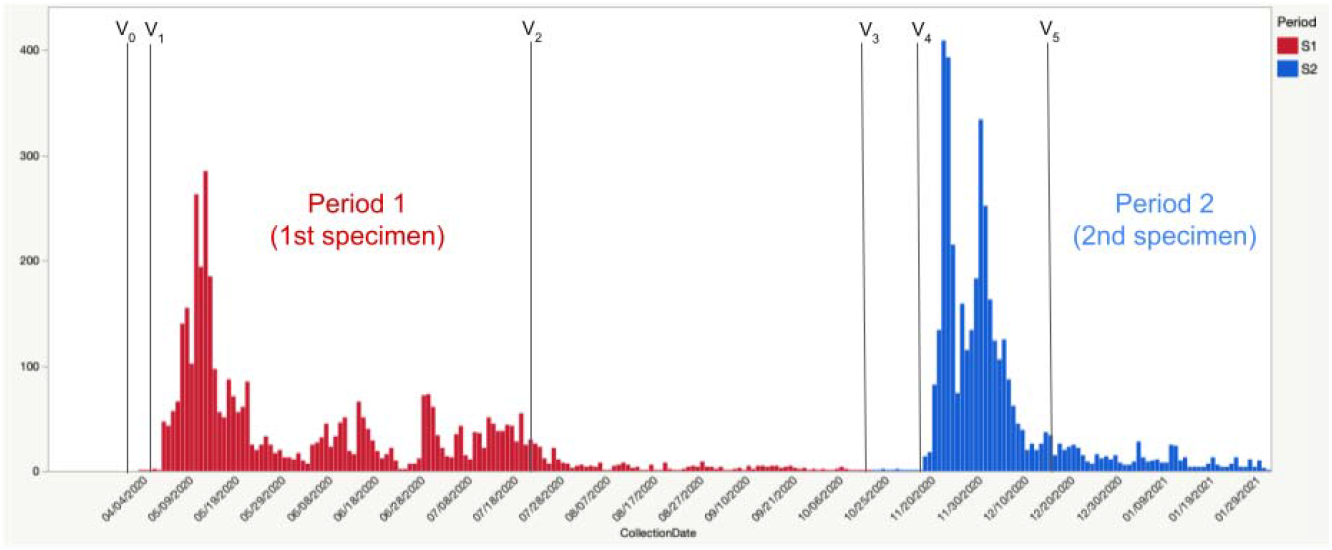
Timing of first (red) and second (blue) dried blood spot specimen collection in the CHASING COVID Cohort Study, including follow-up interview milestones

## Notes

**CONFLICTS OF INTEREST:** None declared

### Competing Interest Statement

The authors have declared no competing interest.

### Clinical Trial

Not an intervention study

### Clinical Protocols

https://bmjopen.bmj.com/content/11/9/e048778

### Author Declarations

The study protocol was approved by the Institutional Review Board at the City University of New York (CUNY) Graduate School for Public Health and Health Policy.

